# Killip-Kimball Classification Validation, Outcomes and Demographics in an Octogenarian Cohort with Acute Coronary Syndrome in a Developing Country: A Third-Level Multicenter 11-Year Experience

**DOI:** 10.1101/2020.09.14.20194480

**Authors:** Diego Ramonfaur, David Eugenio Hinojosa-González, José G. Paredes-Vázquez

**Affiliations:** Harvard Medical School, 25 Shattuck St, Boston, MA 02115, United States; Tecnológico de Monterrey, Escuela de Medicina y Ciencias de la Salud. Dr. Ignacio Morones Prieto O 3000 Monterrey, Nuevo León 64000, México.

**Keywords:** Killip-Kimball, acute coronary syndrome, validation, octogenarian

## Abstract

**Introduction:** The Killip-Kimball Classification (KC) is used to group patients with acute coronary syndrome (ACS) based on their clinical profile. It has proven to be useful while predicting both short- and long-term mortality. Contemporary data in the elderly population is limited. We sought to analyze trends in outcomes of patients 80 years or older admitted for ACS, by Killip Class. In addition, we assess the validity of the KC in this population.

**Methodology:** A retrospective analysis of patients who underwent a catheterization procedure for ACS was performed. ACS was defined as per AHA guidelines, and included STEMI, non-STEMI and Unstable Angina. We determined factors influencing the KC in which patients present to the emergency room. Likewise, we compared inhospital mortality, length of stay, and other outcomes dividing the patients by KC.

**Results:** A total of 133 patients were analyzed. Included were: 86, 9, 23 and 15 patients in KC-I through IV respectively with a mean age of 83. The main comorbidities were hypertension (73%), and diabetes (43%). In-hospital mortality was 12%, which was different between KC groups (p < 0.01). In addition, we found higher KC groups to be associated with acute kidney injury during the hospitalization (p< 0.01).

**Conclusion:** Despite a strong reduction in mortality for elderly patients with ACS in recent decades, patients presenting with ACS and higher Killip class have a high mortality rate, as described in younger cohorts. The Killip-Kimball classification remains a reliable prognostic tool, with applicability in octogenarian patients.

## 1. Introduction

As the population around the world ages, chronic diseases become more prevalent. In particular, the prevalence of coronary artery disease has grown in the past few decades. (1) Hence, patients in the very elderly population with acute coronary syndrome (ACS) are representing a higher proportion of admissions. Traditionally, octogenarian and nonagenarian patients with ACS were not treated with percutaneous coronary intervention (PCI), given the poor prognosis this diagnosis represented for this population and the high risk it represents to undergo an invasive procedure. For these reasons, for many years the very elderly were excluded from clinical trials in light of standardizing treatment for ACS.(2) However, recent data suggests in-hospital morbidity and mortality do not differ greatly when compared with younger patients. (3) Since its first publication in 1967, by Killip and Kimball, this Classification has been used as a clinical estimate of severity in adults with acute heart failure (AHF) secondary to ACS. The KC classifies patients into four groups following the clinical criteria according to Killip and Kimball in their original paper. KC-I: No signs of congestion, KC-II: S3 and basal rales on auscultation, KC-III: Acute pulmonary edema and KC-IV: Cardiogenic Shock. (4) The clinical relevance of this classification is the short and long-term mortality estimation; several authors have attempted to define. (3,5,6) New scoring systems have emerged, in light to further predict the mortality with greater precision. However, the Killip classification remains an easy-to-use tool that has proven to be reliable. The objective of our study is to validate this classification method in patients 80 years or older of a developing country to facilitate rapid urgent treatment decisions and prognosis without the need of wait laboratory testing results as well as an in-hospital predictor of outcomes.

## 2. Methods

A retrospective analysis including octogenarian and nonagenarian patients admitted for Acute coronary Syndrome (ACS) who underwent cardiac catheterization in two academic third-level centers in Mexico from January 2008 to April 2019 was performed. Approval of the pertained ethics committee was obtained accordingly. Every patient 80 years or older admitted for ACS to any of the two centers was included. Types of ACS were divided into 3 categories: acute ST-elevation myocardial infarction (STEMI), non-ST-elevation myocardial infarction (non-STEMI) and unstable angina (UA). These diagnoses were assigned based on criteria according to the definitions of American Heart Association guidelines.(7,8) KC class was retrospectively assigned based on the criteria mentioned above. Patients who had a KC assigned on their admission note were automatically classified under such class. The hospital Electronic Medical Record (EMR) was used to extract patient data and classify according to KC. Our primary endpoint is to find the relationship between this classification and mortality, given current medical and interventional management. Our secondary endpoint is to find other variables that have a relation with a higher Killip Class (KC) in our cohort, such as length of stay (LOS)

### 2.1 Statistical Analysis

Statistical Analysis was performed in SPSS v25 (Boston IBM) and R statistical software (version 3.6.1). Kolmogorov-Smirnov (K-S) test was used to assess normality of distribution of continuous variables. Analysis of variance (ANOVA) and T-Student were used for normally distributed variables while Kruskal-Wallis (KW) and Mann-Whitney U were used for nonparametric, continuous variables. Post Hoc testing included Tukey for ANOVA and Dunn’s test for Kruskal-Wallis. We used an alpha of 0.05 to declare statistical significance in all of the tests performed. Confidence intervals (CI) are expressed with 95% reference range.

## 3. Results

A total of 193 participants fulfilled inclusion criteria. After excluding participants with incomplete EMR, 133 patients were analyzed. The population included 54 (40.6%) females and 79 (59.4%) males. Gender was not different between groups (p=.403). Mean and median age were 83 (IQR 4) and was not statistically different between KC groups (p= 0.361).

Fifty-eight (43.6%) had prior history of Diabetes Mellitus Type 2 (DM2), while 96 (72.9%) had prior Hypertension (HTN) history. 21(15.8%) had past medical history of Coronary Artery Bypass Graft (CABG) surgery. Other cardiovascular pathologies such as stroke were present in 14 (10.5%) of cases. No significant differences were found between KC groups of patient’s prior history and comorbidities on admission including DM2, HTN, dyslipidemia, smoking, COPD, CKD, peripheral vascular disease (PVD), atrial fibrillation (AF), prior stroke or CABG. Findings are summarized in Table 1.

Emergency department (ED) medical treatment was homogeneous between KC groups. 77 (42%) patients received P2Y12 antagonists (p= .908), with either clopidogrel, prasugrel or ticagrelor. 98 (74%) received anticoagulation (p= .638) with either activated factor X inhibitors, unfractioned heparin (UFH) or low molecular weight heparin (LMWH). 74 (55%) received statin therapy (p= .598). 35 (26%) patients received beta blockers (BB) of any kind (p= .325). 98 (74%) patients received aspirin therapy (p= .633). Although there was no significant difference regarding the treatment in the ED between groups, there was a trend towards a higher proportion of patients in KC-IV who received BB as well as aspirin therapy.

A total of 34 (26%) of patients were diagnosed with STEMI, the remaining 99 patients had a diagnosis of either non-STEMI or UA. For this category, there was a significantly lower proportion of patients with STEMI in the KC-I category. Only 4 (3%) patients underwent CABG during their admission after unsuccessful PCI attempt or unfavorable angiography. A total of 92 (69%) patients had at least 1 stent deployed. Nonetheless, stent placement was not different between groups (p= .863).

### 3.1 Primary endpoint

A total of 16 (12%) patients died during hospitalization. 4 (5%) in KC-I, 1 (11%) in KC-II, 5 (22%) in KC-III and 6 (40%) in KC-IV. Mortality was significantly different among classes (p= 0.002), with subgroup significant differences between KC-I and III (p<0.001) as well as for KC-I and IV (p< 0.001).

### 3.2 Secondary endpoint

A total of 3 patients suffered from stroke during hospitalization, 2 (9%) in the KC-III group and 1 (7%) in the KC-IV group. A total of 4 patients suffered reinfarction in our cohort, 1 (1%) in KC-I, 1 (4%) in KC-III and 2 (13%) in KC-IV. Thirty-three patients were documented to have any degree of acute kidney injury (AKI), 13 (15%) in KC-I, 1 (11%) in KC-II, 11 (48%) in KC-III and 8 (53%) in KC-IV. While no differences were found between reinfarction rates and stroke between KC groups, significant differences were found in intra-hospital AKI between groups. (p=0.01). AKI was significant between KC-I and III (p <0.01), as well as KC-I and IV (p <0.01). Even when adjusting development of Acute Kidney Injury in patients without prior Renal comorbidities, differences in Acute Kidney Injury rate was statistically significant between groups (p= 0.001). Median length of stay was 4 days (IQR 4.25). KW test showed no statistically significant difference in LOS between Killip Class groups *(p* = 0.062). However, after adjusting for mortality, differences between KC groups were statistically significant *(p* = 0.007), with differences found between KC-I and K-IV (p<0.05).

### 3.3 Patient History

No significant differences were found between KC groups of patient’s prior history and comorbidities on admission including DM2, HTN, dyslipidemia, smoking, COPD, CKD, peripheral vascular disease (PVD), atrial fibrillation (AF), prior stroke or CABG. Findings are summarized in Table 1

**Table 1.**

|  | KC-I | KC-II | KC-III | KC-IV | *n* | p-value |
| --- | --- | --- | --- | --- | --- | --- |
| DM2 | 39 (45%) | 2 (22%) | 10 (44%) | 7 (47%) | 58 (44%) | .607 |
| HTN | 62 (72%) | 8 (89%) | 17 (74%) | 10 (67%) | 97 (73%) | .634 |
| Smoking | 25 (29%) | 3 (33%) | 8 (24.8%) | 4 (27%) | 40 | .938 |
| Dyslipidemia | 34(40%) | 6 (67%) | 7 (30%) | 3 (20%) | 50 | .116 |
| PVD | 17 (20%) | 1 (11%) | 4 (17%) | 3 (20%) | 25 | .931 |
| AF | 15 (17%) | 2 (22%) | 6 (26%) | 1 (7%) | 24 | .485 |
| Stroke | 6 (7%) | 3 (33%) | 2 (9%) | 3 (20%) | 14 (11%) | .054 |
| CABG | 17 (20%) | 1 (11%) | 3 (13%) | 0 (0%) | 21 (16%) | .249 |
| COPD | 11 (13%) | 1 (11%) | 1 (4%) | 2 (13%) | 15 | .707 |
| CKD | 8 (9%) | 2 (22%) | 4 (17%) | 1 (7%) | 15 | .459 |

### 3.4 Admission Laboratory Values and hemodynamic parameters

Median creatine phosphokinase (CPK) was 102.5 mg/dl. Significant differences were found between groups in CPK values *(p* < 0.001, CI: 231–629) with differences present between groups KC-I and III (p= .001), and KC-I and IV (p = .006). Median creatine phosphokinase muscle-brain (CK-MB) was 24 mg/dl, with significant difference between groups *(p* < .01, CI: 33-68). Particularly between KC-I and IV (p= .013). Hemoglobin (Hb) had a Median value of 12.4 mg/dl, with no differences found between groups *(p*=.442, CI: 11.69-12.76). Median glucose on admission was 138.5 with significant difference between groups *(p* = .002, CI: 151–192). Particularly between KC-II and III (p = .021) and KC-II and IV (p= .021). Median creatinine was 1.81mg/dl, with significance between groups *(p*= .000, CI: 1.13–1.48) and with differences present between KC-I and KC-IV (p= .001) and KC-II and KC-IV (p= 0.004). Mean leucocyte count was 10,382/μL with statistical significance between groups (p= .000). Differences between KC-I vs III (p= .009), and KC-I vs IV (p= .000) were significant.

As far as hemodynamic parameters, admission left ventricular ejection fraction (LVEF), Heart Rate (HR), Systolic (SBP) and Diastolic (DBP) blood pressures were analyzed for each group. Median HR was 85 bpm, significantly different between KC groups. Subgroup analysis revealed statistical significance between KC-I and III (p=.001) as well as KCI-IV (p< .001). As for SBP with a mean of 119.28 mmHg, groups were statistically different (p< .001, CI: 113–125). In subgroup analysis, there were significant differences between KC-I and IV (p< .001). Mean DBP was 70 mmHg, significantly different among groups (p< 0.001, CI: 66–74). In subgroup analysis KC-I and IV (p= .000), as well as KC-III and IV (p= .006) were statistically different. Median LVEF was 50%, with a significant difference between groups (p= .005). In subgroup analysis, KC-I and III were significantly different (p= .026) as well as KC-I and IV (p= .008). Admission laboratory values and hemodynamic parameters for each KC group are enlisted in Table 2.

**Table 2.**

| Parameter | units | KC-I | KC-II | KC-III | KC-IV | p-value |
| --- | --- | --- | --- | --- | --- | --- |
| CPK* | mg/dl | 86(IQR 161) CI<br>(68.2-429.4) | 122 (IQR 817.25)<br>CI (135.2-922.2) | 100(IQR 856.5) CI<br>(13.9-1479.1) | 597.5 (IQR 684) CI<br>(195.5-1090.84) | < .001 |
| CK-MB* | mg/dl | 21.80 (IQR 23.2) CI<br>(19.90-39.63) | 33.05 (IQR 83.3)<br>CI (20.4-134.2) | 23 (IQR 81.7) CI<br>(24.3-134.6) | 35 (IQR 73.8) CI<br>(7.9-152.3) | < .01 |
| Hb* | mg/dl | 12.2 (IQR 3.1) CI<br>(11.03-12.8) | 12 (IQR 0.47) CI<br>(11.53-12.4) | 12.50(IQR 2.45) CI<br>(11.7-13.5) | 12.9 (IQR 2.32) CI<br>(11.9-13.6) | .442 |
| *Leucocytes*^ | 10<sup>3</sup>/μL | 9271 (IQR 4100) CI<br>(8139-10403) | 9383 (IQR 4100)<br>CI (8139-10402) | 11529 (IQR 6350)<br>CI (9391-13668) | 13058 (IQR 7375)<br>CI (10221-15860) | < .001 |
| Glucose* | mg/dl | 140 (IQR 68) CI<br>(142-198) | 102 (IQR 82.7) CI<br>(31.7-187.8) | 159 (IQR135) CI<br>(140.6-247) | 165 (IRQ110.5) CI<br>(135-223.3) | < .01 |
| Creatinine* | mg/dl | 1.0 (IQR 0.5) CI<br>(0.97-1.34) | 0.95 (IQR 45) CI<br>(0.68-1.35) | 1.2 (IQR 0.6) (CI.97-1.61) | 1.86 (IQR.97) CI<br>(1.24-2.77) | < .001 |
| *SBP*^ | mmHg | 113 (IQR 33) (CI<br>116-129) | 115 IQR (20)<br>(CI99-130) | 131 (IQR 44)<br>(CI114-148) | 93 (IQR 43) CI (78-109) | < .001 |
| *DBP*^ | mmHg | 73.3(IQR 23) CI<br>(68.2-78.4) | 62.5 (IQR 14.8) CI<br>(50.4-74.6) | 73.3(IQR 28) CI<br>(64.9-81.7) | 61.3(17.5) CI (53.6-68.9) | < .001 |
| HR* | bpm | 80 IQR (21)<br>CI (73.8-83.4) | 82.5 (IQR 34.5)<br>CI (57.8-96.5) | 94(IQR 37) (CI 81.6-114.7) | 110 (IQR 60) (CI<br>73.3-118.9) | < .001 |
| LVEF* | % | 55(IQR 15) CI (47) | 50(IQR 18) CI (33-64) | 40 (IQR 30) CI (30-46) | 30 (IQR 10) CI (24-40) | .005 |

\*Medians
^Means

### 3.5 Electrocardiographic Characteristics in the emergency department

Admission electrocardiograms were interpreted for abnormalities. Statistically significant differences were found between classes in AV Blocks and ST segment elevations (p=.012) and (p= .001), respectively. Analysis of subgroups revealed statistically significant differences between KC-1 and III (p= .001) in ST Segment Elevations while AV Block was statistically significant between KC-I and III (p= .003) as well as for KC-III and IV (p= .003). Findings of all EKG characteristics are summarized in Table 3.

**Table 3.**

|  | KC1 | KC2 | KC3 | KC4 | *n* | p- value |
| --- | --- | --- | --- | --- | --- | --- |
| Sinus Rhythm | 45 (52%) | 5 (56%) | 18 (82%) | 9 (60%) | 77 (58%) | .097 |
| ST Elevation | 12 (14%) | 4 (44%) | 12 (52%) | 6 (40%) | 34 (26%) | .001 |
| TWA | 40 (47%) | 2 (22%) | 8 (35%) | 4 (27%) | 54 (41%) | .258 |
| AV Block | 21 (25%) | 4 (44%) | 4 (17%) | 11 (73%) | 49 (37%) | .001 |
| LBBB | 13 (15%) | 2 (22%) | 3 (13%) | 3 (20%) | 21 (16%) | .892 |

Sample size (proportion)

## 4. Discussion

The prognosis of the very elderly after ACS has improved in part due to refinement of treatment and PCI protocols. (9–11) A recent metanalysis that included 32 studies corroborated the importance of prioritizing PCI in patients admitted for STEMI, where they found an Odds Ratio of 1.52 of short-term mortality in patients with > 90 minutes of door-balloon time compared to those with < 90 minutes of door-balloon time. (12) These results included but were not limited to elderly patients. Amongst causes of death in the very elderly population with ACS, cardiogenic shock and acute renal failure are most important culprits reported. (13) Despite the Killip and Kimball classification being an old and simple scoring system, research around this classification has prevailed and many centers continue to use it for clinical profile classification purposes. A landmark contribution made in 2019 by Zadok and collaborators demonstrated statistical significance in both short- and long-term mortality in patients with higher KC. In their cohort’s large sample size, the added value of following patients for a longer period of time demonstrated that higher KC was associated with an increased mortality rate at 1-year. (5) This finding is an important contribution since KC has been traditionally used in the acute setting and not necessarily meant as a long-term mortality predictor. Consistently with this publication, a study done by DeGeare and collaborators in 2001 found, in a large cohort, a significant relationship between Killip Class and 6-month mortality, although excluding participants in class IV. Our study had consistent results with the literature that studies a wider population in developed countries. Our results, based on a multicenter cohort in a developing country, found robust evidence towards the value of utilizing the Killip-Kimball classification in the setting of ACS in patients 80 years or older. In concordance with recent data, this study strengthens the prognostic value, as well as the clinical implications of the routine implementation of this classification. The overall mortality in our population was 8% before excluding patients with incomplete EMR, after which the mortality rose to a more overwhelming 12%. Hence, the results obtained from this study in octogenarians and nonagenarians, shows very similar mortality rates within each clinical profile compared to studies with younger cohorts. (3) Our findings pertaining hemodynamic measurements were as expected. By definition, KC-IV must have shock, which almost invariably comes with tachycardia and decreased SBP and DBP, as worsening cardiac contractility and activation of compensatory mechanisms subside. (14,15) Nonetheless, we analyzed these results to provide insight on the degree of hemodynamic compromise as well as to have LVEF as another point of reference. Higher KC tended towards higher heart rate.

These findings further solidify the decision to aggressively treat elderly patients with ACS, rather than utilizing a conservative approach as well as determine a more reliable prognosis after a interventional procedure. (16,17) Our study strengthens the growing literature supporting the use of KC as an in-hospital mortality prognostic indicator. We also describe how this classification relates to other paraclinical studies and laboratory values. In spite of a population of a developing country, our patients had an acceptable hemoglobin concentration amongst all groups. This reflects an overall wellness status within the cohort. Global renal function was reduced, with a median creatinine of 1.81 mg/dl. This rather expected finding, thus partially explained by the acute ACS insult, also reflects the important association between renal and cardiovascular disease.

Using all this information in conjunction may give a much clearer picture and expectation of the very elderly patient’s clinical scenario and can validate the use of the KC in this special group of patients.

### 4.1 Limitations

Since this is a retrospective chart review, there are potential confounders that could not have been corrected for or taken into account. Our database consisted of patients who underwent cardiac catheterization. As a result, patients deceased at the emergency department were not included in this cohort, this limitation has potential implications on underestimating the severity of ACS in our population. The treatment patients received was at the discretion of the attending cardiologist, making it a heterogeneous cohort in terms of management. Because of the third-level nature of our two centers, there is limited generalizability towards the whole octogenarian and nonagenarian community, depending on the accessibility to healthcare facilities.

## 5. Conclusion

In recent decades, the population has transitioned towards an older demographic. As research in cardiovascular disease continues to gain interest and popularity, newer and better therapies emerge. This inevitably will lead to increased prevalence of cardiovascular diseases, as the mortality rate decreases. Our contribution aims to emphasize the prompt and aggressive treatment for elderly patients with ACS and unfavorable clinical profile. We add evidence to the limited literature describing short term mortality in octogenarian patients with ACS. The KC has good prognostic value for the very elderly population and has proven a good tool to reliably stratify patients according to their clinical profile. These new insights to the use of the Killip-Kimball classification in the very elderly population should help clinicians have a better sense of the outcomes of octogenarian patients with ACS in the emergency department. We encourage new initiatives to further polish the prognostic value of this classification in the current era with controlled studies to define better and more personalized treatment strategies for this growing population.

## Data Availability

Dataset may be provided upon request

